# Out-of-pocket costs and productivity losses associated with influenza illness and caregiving in a household study in the United States, 2024–2025

**DOI:** 10.64898/2026.09.10.26362662

**Authors:** Elizabeth B. White, Jamison Pike, Celibell Y. Vargas, Yuwei Zhu, Caroline A. O’Neil, Jessica E. Biddle, Lydia Bristol, Emily McNair, Son H. McLaren, Ellen Sano, Theresa A. Scott, Rachel Presti, Stephanie A. Fritz, Lisa A. Prosser, Melissa S. Stockwell, Carlos G. Grijalva, Jennie H. Kwon, Alexandra M. Mellis

## Abstract

**Background:** Non-medically attended (NMA) and outpatient medically attended (MA) influenza illnesses contribute substantially to influenza disease burden in the United States. We leveraged a household study to estimate out-of-pocket and lost productivity costs of NMA and MA influenza illness.

**Methods:** The Respiratory Virus Transmission Network for Influenza was a 2024–2025 case-ascertained household study at 3 U.S. sites. Influenza-positive index patients and household contacts self-reported daily symptoms, over-the-counter (OTC) medication use, medical care-seeking, time missed from work or activities, and self-collected nasal swabs for PCR testing. We included participants with PCR-confirmed influenza infection and influenza-like-illness symptoms who ever (MA) or never (NMA) sought medical care during follow-up. We obtained costs of OTC medications from retail websites and productivity costs from published values. We calculated OTC medication and lost productivity costs in 2025 USD for MA and NMA influenza separately and household-level lost productivity costs among caregivers for MA and NMA household members.

**Results:** Median out-of-pocket OTC medication costs were $6.38 (interquartile range [IQR] $2.92–$11.60) among 594 MA and $3.72 (IQR $1.50–$7.05) among 153 NMA influenza illnesses. Among participants ≥15 years of age, median lost productivity was $315 ($149–$743) for MA and $248 ($0–$624) for NMA illnesses. Median household-level lost productivity for caregiving was $148 (households with 1 ill child), $379 (multiple ill children), and $0 (≥1 ill adult).

**Conclusions:** Influenza causes substantial lost productivity costs within households. These estimates can inform future economic burden and cost effectiveness analyses.

## Introduction

Seasonal influenza epidemics in the United States cause not only a substantial burden of illness, hospitalization, and death (1), but also large economic impacts for individuals and society. The economic burden of influenza in the United States has been estimated at approximately $11.2 billion each year, including $3.2 billion in direct costs associated with medical care and $8.0 billion in indirect costs resulting from lost productivity (2). However, these previous estimates have had to rely on assumptions for several inputs, notably the costs of non-medically attended (NMA) influenza illnesses.

The direct healthcare and indirect costs of medically attended (MA) influenza illness have been estimated previously through surveillance data and interviews (3). However, out-of-pocket costs of over-the-counter (OTC) medications have not been studied for NMA or MA influenza. In addition, the productivity losses and indirect costs associated with NMA influenza illnesses have not been well characterized because these individuals do not seek medical care, may not know of their influenza infection, and are difficult to identify for research studies. Nonetheless, millions of NMA influenza illnesses occur each year and contribute to the overall economic burden of influenza. Productivity losses associated with influenza can arise from time missed from work (market productivity losses) and from lost unpaid labor within the home, volunteering, and school attendance (non-market productivity losses). In addition, caregiving for household members who are ill with influenza can result in productivity loss for the caregiver. Estimating these costs is important to accurately assess the potential economic benefits of influenza prevention measures, since NMA influenza illnesses are common (4, 5). Case-ascertained household transmission studies, in which household contacts of the first known influenza-positive (index) case are prospectively tested each day, are an efficient design to identify both MA and NMA influenza illnesses, characterize caregiving within households, and estimate the associated costs.

The objective of this study was to produce estimates of the costs of MA and NMA influenza illness and associated household-level caregiving costs in the Respiratory Virus Transmission Network Influenza (RVTN-Flu) study during the 2024–2025 season. We aimed to assess out-of-pocket OTC costs and to describe both market and non-market productivity losses among participants with NMA and MA influenza and caregivers. These data are important to understand the financial impact of influenza on millions of people in the United States each year and to produce more accurate estimates of the national economic burden of influenza.

## Methods

### Study design and population

The Respiratory Virus Transmission Network for Influenza (RVTN-Flu) was a case-ascertained household transmission study during the 2024–2025 influenza season conducted at research sites in three U.S. states: Missouri (Washington University in St. Louis School of Medicine), New York (Columbia University Irving Medical Center), and Tennessee (Vanderbilt University Medical Center). RVTN has previously published findings related to influenza illness symptoms (6), transmission dynamics (7, 8), and vaccine effectiveness (9).

### Data collection

Our analysis aimed to estimate out-of-pocket costs and lost productivity costs, including market and non-market lost productivity arising from both ill persons and their caregivers.

In RVTN-Flu, index cases who tested positive for influenza at an outpatient clinic or emergency department and their household contacts were recruited and enrolled within 6 days of index case symptom onset. Index cases who were hospitalized were excluded. After participants or guardians provided written consent, each participant provided demographics and current season influenza vaccination status. Participants completed a retrospective diary of symptoms, medication use, and productivity losses for each previous day between index case symptom onset (or positive test) and enrollment. Following enrollment, daily diaries and nasal swabs were prospectively collected for 7 days. Nasal swabs were self-collected daily and shipped to the central RVTN-Flu laboratory at Vanderbilt University Medical Center for processing and influenza testing using the Hologic Panther Fusion^TM^ quantitative RT-PCR assay (AW-16832).

Daily diary questions regarding medication use and productivity losses are available in full in the **Supplement** and briefly described here. Daily medication use was reported for 12 specific OTC medications or medication categories selected in consultation with clinical partners; we selected medication categories that would be recognizable to participants, and we considered costs for a range of specific name brand and store brand medications within each category (see **Supplementary Materials**). Medication categories were: cold and flu or sinus medicine, Theraflu, Zicam, ibuprofen, acetaminophen, naproxen, aspirin, Mucinex, nasal spray, Sudafed, allergy medicine, or other medication. For each medication, participants responded yes or no to, “On [date] did [name] take [medication];” if they said yes, they were asked how many doses (with possible answer choices 1, 2, or 3+).

Daily productivity losses were reported through 4 questions asking if the participant had missed work, school, or other activities that day due to their own illness or to care for an ill household member. If the participant selected yes to any of those questions, they were prompted to provide the number of hours missed that day. In addition to daily diaries, adult participants were invited to complete a modified Work Productivity and Activity Impairment (WPAI) questionnaire at the 30-day follow-up time point. The WPAI, which typically assesses impairments with a 7-day recall period, is a validated tool used by health economists to estimate productivity losses due to illness (10). Our adapted version is available in the **Supplement**. Briefly, adults were asked a series of questions regarding their employment status, ability to work remotely, availability of paid sick leave, and whether they had missed time from work or activities due to their own illness or to care for an ill household member. If a participant answered “yes” to any question about having missed work or other activities, they were prompted to provide the weekly number of hours missed due to influenza during the 4-week period between study enrollment and 30-day follow up.

### Cost inputs

For out-of-pocket costs associated with OTC medications, we used consumer websites such as pharmacies and online retailers to obtain pre-tax costs per dose for each of the medication categories listed above in November 2025. For “other medication,” we used the average cost per dose across the specifically elicited medications in this study. We estimated costs separately for children aged <12 years and for people aged ≥12 years; 12 years is the age at which OTC medications can generally be taken according to the adult dosing guidelines, and OTC medications formulated for children aged <12 years are priced differently than those for persons ≥12 years. More information about OTC medication cost inputs is provided in the **Supplement**.

For indirect costs resulting from lost market and non-market productivity, we used published values of annual productivity estimated using the US Census Bureau’s American Community Survey (ACS) data in the United States (11) (see **Table S1**). Annual market and non-market productivity values by age group were separately converted to hourly estimates, assuming a 365-day year, a 40-hour workweek, and an 8-hour workday, and to 2025 USD. Consistent with ACS estimates, children aged <15 years were not assumed to contribute directly to economic productivity, and children aged 15–17 were assumed to contribute the same hourly market and non-market productivity as adults aged ≤24 years (**Table S1**).

### Illness cost analysis

To estimate out-of-pocket costs associated with OTC medication use, we used participants reported frequency and duration of use in days and total doses for each of the 12 OTC medications and for OTC medications overall. We used medication-specific recommended dosing to impute the number of daily doses for participants who indicated taking a medication with “3+ doses” and conducted sensitivity analyses assuming exactly 3 doses were taken. We used medication specific dosing instructions and participant demographics to impute the number of doses taken if dosing data was blank or unknown. Total out-of-pocket costs were calculated for each participant by multiplying medication doses by the per-dose cost of each medication and summing these costs across all medications and illness days. Detailed methods are provided in the **Supplementary Materials**.

We estimated out-of-pocket costs and lost productivity due to illness for two groups: participants with NMA influenza illness and those with MA influenza illness. We defined NMA influenza as participants with a PCR-confirmed influenza infection who answered “no” to having sought medical care in all of their daily diaries. We defined MA influenza as participants with a PCR-confirmed influenza infection who sought medical care either as an index participant or during follow-up, as indicated in daily diaries. Consistent with how CDC estimates the national burden of influenza illness, we only included symptomatic NMA and MA influenza illnesses with self-reported influenza-like illness (defined as fever or chills plus cough or sore throat).

To estimate productivity losses due to illness, we used daily diary reports of hours missed from work, school, or activities. For children aged <15 years, we summarized hours missed from school or activities. For participants aged ≥15 years, we assumed that “hours missed from work or school” referred to “work,” or paid employment. Since daily diaries prompted participants to consider an 8-hour workday, we converted any daily reports of >8 hours missed to 8. We summed each participant’s hours missed from work due to illness across diary days and multiplied by the age-group specific hourly market productivity value for adults. For non-market productivity losses among adults, we summed each participant’s hours missed from other activities due to illness across all diaries days and multiplied by the age group specific hourly non-market productivity value.

In order to understand data completeness and utility of the validated WPAI tool administered at 30-day follow-up, we also estimated productivity losses due to illness among adults who responded to the WPAI. We also summarized data not collected through daily diaries, including frequencies of employment, ability to work remotely when ill, availability of paid sick leave, and hours missed from work and activities due to illness by adults with MA or NMA influenza who completed the WPAI. When hours missed from work or activities were left blank, we assumed they were 0.

### Caregiving cost analysis

Due to the study design, we were unable to attribute specific caregiving hours to specific household members and therefore performed a combined analysis for MA and NMA influenza. To estimate household-level productivity losses due to caregiving, we summed the total number of hours that participants aged ≥15 reported missing work or activities to provide care for their ill household member(s) and multiplied by the corresponding age-group specific market (for missed work) or non-market (for missed activities) hourly productivity value (**Table S1**). To examine caregiving needs of a household by age of the ill person(s), we grouped households where influenza illnesses occurred into 1) children aged <18 years only, 2) children aged 0–4 years only (a subset of group 1), 3) children aged 5–17 years only (a subset of group 1), 4) adults aged ≥18 years only, and 5) all ages (all households). Within each group, we summarized caregiving hours and costs separately for households with only 1 influenza illness and those with >1 influenza illness.

For all analyses, we report medians and interquartile ranges (IQRs) by age group and overall. In addition, we report means and 95% confidence intervals (CIs) in the **Supplement**. Bias-Corrected and Accelerated CIs were obtained by bootstrapping with 1,000 resamples using the “boot.ci” function in the *boot* package in R.

### Ethical considerations

The study protocol was reviewed by the Washington University in St. Louis School of Medicine’s Institutional Review Board with reliance from participating sites and by CDC and was conducted consistent with applicable federal law and CDC policy (45 CFR 46.102(l)(2)). All participants or their guardians provided written consent. All analyses were conducted using R version 4.5.3.

## Results

### Study population of NMA illnesses

During the 2024–25 RVTN-Flu study, 1,591 participants were enrolled, of whom 594 (37%) had a MA influenza illness and 153 (10%) met the definition of NMA influenza illness. Compared with all RVTN-Flu 2024–25 participants, those with MA influenza tended to be younger (<18 years), and those with NMA influenza were more likely to have been enrolled at the New York study site and be unvaccinated for influenza in the current season (**Table 1**).

**Table 1.**
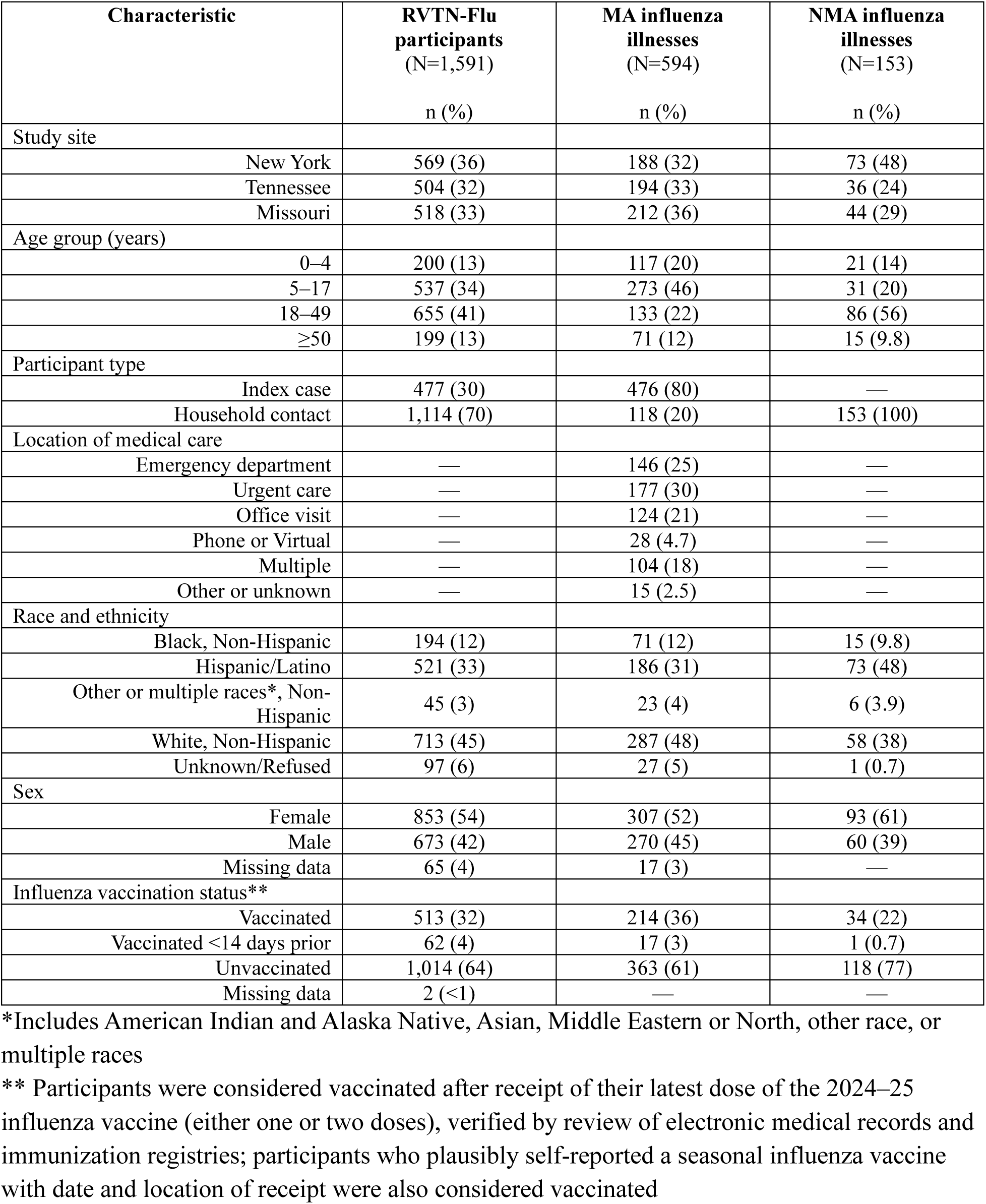
Characteristics of all RVTN-Flu 2024-25 participants, those with MA influenza illness, and those with NMA influenza illness.

| Characteristic | RVTN-Flu participants<br>(N=1,591) | MA influenza illnesses<br>(N=594) | NMA influenza illnesses<br>(N=153) |
| --- | --- | --- | --- |
|  | n (%) | n (%) | n (%) |
| Study site |  |  |  |
| New York | 569 (36) | 188 (32) | 73 (48) |
| Tennessee | 504 (32) | 194 (33) | 36 (24) |
| Missouri | 518 (33) | 212 (36) | 44 (29) |
| Age group (years) |  |  |  |
| 0–4 | 200 (13) | 117 (20) | 21 (14) |
| 5–17 | 537 (34) | 273 (46) | 31 (20) |
| 18–49 | 655 (41) | 133 (22) | 86 (56) |
| ≥50 | 199 (13) | 71 (12) | 15 (9.8) |
| Participant type |  |  |  |
| Index case | 477 (30) | 476 (80) | — |
| Household contact | 1,114 (70) | 118 (20) | 153 (100) |
| Location of medical care |  |  |  |
| Emergency department | — | 146 (25) | — |
| Urgent care | — | 177 (30) | — |
| Office visit | — | 124 (21) | — |
| Phone or Virtual | — | 28 (4.7) | — |
| Multiple | — | 104 (18) | — |
| Other or unknown | — | 15 (2.5) | — |
| Race and ethnicity |  |  |  |
| Black, Non-Hispanic | 194 (12) | 71 (12) | 15 (9.8) |
| Hispanic/Latino | 521 (33) | 186 (31) | 73 (48) |
| Other or multiple races*, Non-Hispanic | 45 (3) | 23 (4) | 6 (3.9) |
| White, Non-Hispanic | 713 (45) | 287 (48) | 58 (38) |
| Unknown/Refused | 97 (6) | 27 (5) | 1 (0.7) |
| Sex |  |  |  |
| Female | 853 (54) | 307 (52) | 93 (61) |
| Male | 673 (42) | 270 (45) | 60 (39) |
| Missing data | 65 (4) | 17 (3) | — |
| Influenza vaccination status** |  |  |  |
| Vaccinated | 513 (32) | 214 (36) | 34 (22) |
| Vaccinated <14 days prior | 62 (4) | 17 (3) | 1 (0.7) |
| Unvaccinated | 1,014 (64) | 363 (61) | 118 (77) |
| Missing data | 2 (<1) | — | — |

\*Includes American Indian and Alaska Native, Asian, Middle Eastern or North, other race, or multiple races
\*\* Participants were considered vaccinated after receipt of their latest dose of the 2024–25 influenza vaccine (either one or two doses), verified by review of electronic medical records and immunization registries; participants who plausibly self-reported a seasonal influenza vaccine with date and location of receipt were also considered vaccinated

### OTC medication usage and out-of-pocket costs

At any time during follow-up, 572 (96%) and 137 (90%) participants with MA and NMA influenza, respectively, took OTC medication for their illness (**Table 2**). Participants with MA influenza took a median of 12 (6–20) medication doses over a median of 5 (3–7) days, while those with NMA influenza took a median of 7 (IQR 4–14) doses over a median of 3 (IQR 2–5) days. For MA influenza, the overall median cost of OTC medications per participant was $6.46 (IQR $3.13–$11.88). For NMA influenza, the overall median cost of OTC medications per participant was $3.72 (IQR $1.50–$7.56). Within NMA and MA illness groups, results were similar for children aged <12 years and for participants aged ≥12 years. Frequency, durations, and doses of specific medication categories are reported in the Supplement; the most commonly taken OTC medications were similar across NMA and MA illness groups and included cold and flu medication (among those aged ≥12 years), acetaminophen (all ages), and ibuprofen (all ages). Medication-specific usage and costs, costs from the minimum dose sensitivity analysis, and means and bootstrapped 95% CIs are also presented in **Tables S2–S5**.

**Table 2.** Percentage of participants with MA or NMA influenza illness who ever took OTC medications, duration of medication use, number of doses taken, and OTC medications costs (in 2025 USD) per illness.

|  | **MA influenza (N=594)** | **NMA influenza (N=153)** |
| --- | --- | --- |
| **Took OTC medication, n (%)** |  |  |
| Overall | 572 (96) | 137 (90) |
| <12 years | 306 (98) | 36 (95) |
| ≥12 years | 266 (95) | 101 (88) |
| **Medication days, median (IQR)** |  |  |
| Overall | 5 (3–7) | 3 (2–5) |
| <12 years | 5 (3–6) | 3 (1.25–5) |
| ≥12 years | 5 (4–7) | 3 (2–5) |
| **Medication doses, median (IQR)** |  |  |
| Overall | 12 (6–20) | 7 (4–14) |
| <12 years | 11 (6–18) | 7 (4.25–11.5) |
| ≥12 years | 13 (6–23) | 8 (3–14.5) |
| **Medication cost, median (IQR)** |  |  |
| Overall | 6.38 (2.92–11.60) | 3.72 (1.50–7.05) |
| <12 years | 6.50 (4.06–11.07) | 4.60 (2.32–7.48) |
| ≥12 years | 6.03 (2.00–13.12) | 3.20 (1.10–7.05) |

### Productivity losses and costs due to illness

Most participants (512/594 [86%] with MA influenza and 108/153 [71%] with NMA influenza) reported ever missing work, school, or other activities due to their illness. However, 82/594 (14%) participants with MA and 45/153 (29%) with NMA influenza reported 0 hours of lost productivity, resulting in skewed data. Means and bootstrapped 95% CIs are provided in **Table S6.** The median missed time from work, school, or activities due to influenza illness was 32 hours (4 work or school days) for participants with MA influenza and 16 hours (2 days) for participants with NMA influenza. Among participants aged ≥18 years, the median productivity loss was $256 (market) and $41 (non-market) per MA influenza illness and $210 (market) and $41 (non-market) per NMA influenza illness.

In the analysis of WPAI data, 186 adults with MA influenza completed the WPAI and reported productivity hours lost due to illness; participants reported a median (IQR) of 24 (16–36) hours of missed work and 20 (9–39) hours of missed activities while ill with influenza. Remote work and paid sick leave were available to 43% and 67%, respectively, of 150 employed participants with MA influenza. Among 86 adults with NMA influenza who completed the WPAI and reported productivity losses due to illness, influenza led to a median of 0 (0–11) hours of missed work and 0 (0–0) hours of missed activities. Remote work was available to 44% and paid sick leave to 58% of 55 employed participants with NMA influenza who completed the WPAI.

### Productivity losses and costs due to caregiving

Among all households with ≥1 member with MA or NMA influenza, ≥1 person aged ≥15 years reported missing time from work or activities to care for an ill household member in 117 (41%) households with 1 influenza illness and in 122 (62%) households with >1 influenza illness. The median (IQR) hours of work or activities missed by caregivers in households with 1 influenza illness were 0 (0–16), corresponding to $0 ($0–$356) in lost productivity; in households with >1 influenza illness, caregiver productivity losses were 12 (0–32) hours and $238 ($0–$674). For households with all influenza illnesses occurring in children aged <18 years, the median (IQR) caregiver productivity loss was 8 (0–27) hours and $148 ($0–$504) when there was 1 influenza illness in the household, and 16.5 (0–39.5) hours and $379 ($12–$818) in households with >1 influenza illness. In households where all influenza illnesses occurred among adults, the median caregiver productivity losses were 0 hours and $0. Full results are provided in **Table 4**. Similar to productivity losses due to illness, many households reported 0 caregiving hours resulting in skewed data. Means and 95% CIs are reported in **Table S7**.

**Table 3.** Median (IQR) lost productivity hours and costs (in 2025 USD) due to illness, by age group and overall, among 747 participants with influenza illness.

| Age group | N ill | Hours missed from work/school | Market productivity loss (cost) | Hours missed from activities | Non-market productivity loss (cost) | Total hours missed | Total productivity loss (cost) |
| --- | --- | --- | --- | --- | --- | --- | --- |
| *Medically attended influenza* |  |  |  |  |  |  |  |
| 0–4 | 117 | 16 (0–32) | — | 7 (0–24) | — | 22 (0–49) | — |
| 5–14 | 242 | 24 (16–33) | — | 12 (2–4) | — | 36 (24–53) | — |
| 15–17 | 31 | 24 (16–32) | 111 (74–148) | 8 (2–19) | 37 (9–89) | 36 (24–48) | 166 (111–222) |
| 18–49 | 133 | 16 (8–32) | 297 (149–647) | 4 (0–12) | 33 (0–126) | 26 (9–40) | 416 (210–787) |
| $\geq 50$ | 71 | 21 (5–32) | 357 (22–714) | 7 (0–20) | 62 (0–178) | 30 (15–48) | 455 (128–776) |
| All ages | 594 | 22.5 (8–32) | 256 (92–630) | 8 (0–20) | 41 (0–126) | 32 (16–48) | 315 (149–743) |
| *Non-medically attended influenza* |  |  |  |  |  |  |  |
| 0–4 | 21 | 8 (0–24) | — | 1 (0–8) | — | 16 (0–27) | — |
| 5–14 | 24 | 16 (2–24) | — | 6 (1–17) | — | 22 (6–37) | — |
| 15–17 | 7 | 12 (0–24) | 55 (0–111) | 8 (0–18) | 37 (0–84) | 19 (0–42) | 88 (0–195) |
| 18–49 | 86 | 8 (0–22) | 210 (0–590) | 4 (0–12) | 43 (0–124) | 16 (0–31) | 297 (0–660) |
| $\geq 50$ | 15 | 12 (0–26) | 256 (0–449) | 1 (0–16) | 8.9 (0–143) | 16 (0–32) | 256 (0–449) |
| All ages | 153 | 8 (0–23) | 210 (0–541) | 3 (0–9) | 41 (0–123) | 16 (0–32) | 248 (0–624) |

**Table 4.** Frequency of caregiving, duration of household ILI and caregiving, and median (IQR), market, non-market, and total productive hours missed and lost costs (2025 USD) due to caregiving, by age group of ill household member(s)

| Age of influenza case(s) in HH* | N HH | ILI days in HH | Any care-giving, n (%) | Caregiving days in HH | Hours missed from work by caregiver(s) | Market productivity loss of caregiver(s) (dollars) | Hours missed from activities by caregiver(s) | Non-market productivity loss of caregiver(s) (dollars) | Total hours missed by caregiver(s) | Total productivity loss of caregiver(s) (dollars) |
| --- | --- | --- | --- | --- | --- | --- | --- | --- | --- | --- |
| *Children only (aged <18 years)* |  |  |  |  |  |  |  |  |  |  |
| 1 flu | 190 | 2 (1–4) | 103 (54) | 1 (0–3) | 4 (0–16) | 92 (0–402) | 0 (0–8) | 0 (0–112) | 8 (0–27) | 148 (0–504) |
| >1 flu | 44 | 4 (2–5) | 30 (68) | 2 (0–4) | 10.5 (0–24) | 290 (0–630) | 3.5 (0–8.5) | 55 (0–142) | 16.5 (0–39.5) | 379 (12–818) |
| *Children only (<5 years)* |  |  |  |  |  |  |  |  |  |  |
| 1 flu | 49 | 2 (0–4) | 29 (59) | 1 (0–3) | 8 (0–16) | 120 (0–297) | 3 (0–8) | 55 (0–112) | 9 (0–24) | 208 (0–386) |
| >1 flu | 6 | 3.5 (2–4) | 4 (67) | 2.5 (0–7) | 9 (0–44) | 193 (0–1,040) | 6.5 (0–16) | 83 (0–216) | 21 (0–61) | 356 (0–1,096) |
| *Children only (5–17 years)* |  |  |  |  |  |  |  |  |  |  |
| 1 flu | 141 | 2 (1–4) | 74 (52) | 1 (0–3) | 2 (0–16) | 73.76 (0.00, 419.84) | 0 (0–8) | 0.00 (0.00, 122.40) | 4 (0–28) | 112.36 (0.00, 523.86) |
| >1 flu | 20 | 4 (2.5–5.5) | 16 (80) | 2 (1–4) | 14 (6–24) | 367 (131–630) | 5 (0–8) | 78 (5–124) | 19 (7–35) | 437 (154–818) |
| *Adults only* |  |  |  |  |  |  |  |  |  |  |
| 1 flu | 95 | 3 (1–4) | 14 (15) | 0 (0–0) | 0 (0–0) | 0 (0–0) | 0 (0–0) | 0 (0–0) | 0 (0–0) | 0 (0–0) |
| >1 flu | 34 | 4 (3–6) | 11 (32) | 0 (0–1) | 0 (0–4) | 0 (0–32) | 0 (0–0) | 0 (0–0) | 0 (0–4) | 0 (0–52) |
| *Any age* |  |  |  |  |  |  |  |  |  |  |
| 1 flu | 285 | 2 (1–4) | 117 (41) | 0 (0–2) | 0 (0–12) | 0 (0–238) | 0 (0–6) | 0 (0–78) | 0 (0–16) | 0 (0–356) |
| >1 flu | 192 | 4 (3–6) | 122 (64) | 1 (0–3) | 8 (0–24) | 149 (0–539) | 2 (0–15.5) | 25 (0–145) | 12 (0–32) | 238 (0–674) |

\*Age group definitions: “Children aged 0–4 years only” and “Children aged 5–17 years only” are subsets of “Children only (aged <18 years).” “Any age” includes “Children only (aged <18 years),” “Adults only,” plus all other households with a mix of children and adults with influenza.

## Discussion

In this household study conducted during the 2024–2025 influenza season in the United States, we found that influenza illnesses were associated with median per-illness out-of-pocket OTC medication costs of $6.38 for MA influenza and $3.72 in NMA influenza. Each illness in a person aged ≥15 years resulted in a median of $315 (MA influenza) and $248 (NMA influenza) in productivity losses. We also found substantial household-level productivity losses associated with missed work or activities by a caregiver when influenza illnesses occurred in children, especially when >1 child in the household was ill. Considering that during the 2024–2025 influenza season there were an estimated 51 million illnesses and 23 million outpatient medical visits due to influenza in the United States (12), many of which occurred among children and may have required caregiving within the household, our findings highlight that even non-severe influenza illnesses pose a substantial economic burden on both the individual and national scale.

To our knowledge, our study is among the first to provide real-world estimates of out-of-pocket costs of OTC medications. In previous studies estimating the economic burden of influenza in the United States, out-of-pocket costs associated with OTC medications have been assumed due to a lack of empirical data (2, 13–15). The most recent assumption was a mean of $7 per illness in 2015 USD, corresponding to $9.59 in 2025 USD (2). In this study, we used self-reported medication usage and average retail costs and obtained similar estimates for MA influenza (median $6.38, mean $9.11) but slightly lower estimates NMA influenza (median $3.72, mean $6.03). Having more accurate per-illness costs is especially important when they are used to estimate the economic burden resulting from millions of influenza illnesses occurring in the United States each year.

This analysis found substantial lost productivity costs associated with influenza, including from caregiving. We found that people aged ≥15 years with MA influenza missed a median of 3–4 days, totaling $315, of work, school, and other activities while ill, and those with NMA influenza missed about 2 days, or $248. At the national level, these estimates correspond to millions of lost school and work days and billions of dollars in lost productivity costs during the 2024–25 season. A systematic review of the impact of influenza on work productivity found that, across diverse studies and populations, influenza consistently led to workplace absenteeism and decreased productivity even if working while ill (16). Our findings from the subset of participants who completed the WPAI showed inconsistent access to remote work and paid sick leave while ill. However, due to the small sample size of participants who responded to the WPAI , we were not able to fully characterize these non-monetary impacts of influenza on productivity in this study. In addition, participants generally reported fewer hours of missed work and activities through the WPAI than through daily diaries, especially for NMA illnesses. Further studies using the WPAI are warranted, ideally with a larger study population and as a standalone survey closer to the time of illness to reduce survey fatigue and improve recall.

Our study found that some influenza illnesses, particularly among children, incur substantial lost productivity costs due to caregiving. More than half of households with ≥1 ill child required an adult to miss work or activities to provide care. About one third of the resulting lost productivity costs arose from non-market productivity losses due to missing time from activities besides paid employment. In total, this study found that caregivers in households with ill children aged <5 years incurred $208 in lost productivity costs for one ill child, or $356 if multiple children were ill. These estimates are somewhat higher than in a previous study that used in-depth interviews to ascertain caregiving time, which reported that caregivers of children with influenza aged <5 years treated in outpatient clinics missed a median of 11 work hours or about $127 (converted 2025 USD), while caregivers of children with influenza aged <5 years treated in the emergency department missed 19 hours of work or about $187 (converted to 2025 USD) (3). Our findings suggest the importance of including non-market productivity losses to fully capture the economic impact of caregiving for influenza in a household.

This study has several limitations. First, our study design was unable to estimate the caregiving costs for MA and NMA illnesses separately. To provide these estimates stratified by care seeking, future studies could use a study design that ascertains households with NMA influenza illnesses only or adapt data collection methods to allow attribution of caregiving hours to specific persons in households on days when multiple people are ill. Second, our study was conducted in three states, and our findings might not be representative of all states due to differing taxes on OTC medications. For example, TN is the only state represented in this analysis that taxes OTC medications (9.75% in state and local taxes); our estimates, however, represent pre-tax costs that may underestimate actual out-of-pocket expenses depending on state of residence. Third, our study was not designed to ascertain direct outpatient or emergency department healthcare costs of MA influenza illnesses, or any of the direct or indirect costs of influenza hospitalizations; these costs have been reported elsewhere (3, 17). Fourth, our adult study population included mostly working-age adults, and therefore our results do not generalize to older adults. Finally, we used daily diaries, rather than the validated WPAI, for our productivity loss estimates due to the higher response rate and daily reporting rather than relying on 30-day recall. Our findings might differ from those of other studies using the WPAI alone.

## Conclusion

In this study during the 2024–25 influenza season, we found that MA and NMA influenza illnesses were associated with <$10 in out-of-pocket costs and approximately $315 (MA) and $248 (NMA) in lost productivity costs per illness. Caregivers experienced substantial productivity losses in households with ≥1 child ill with influenza. These estimates are among the first for non-medically attended influenza and can be used in future economic burden and cost effectiveness analyses. Additional studies to characterize lost productivity would provide further insight into the impacts of influenza on households and the workforce in the United States beyond monetary losses.

## Supporting information

Supplemental methods and results

Text of questionnaires

## Data Availability

The data underlying this article will be shared on reasonable request to the corresponding author.

## Acknowledgments

Washington University in St. Louis: Kim T. Vu, Regine Burton, Francesca Yerbic, Lucy Vogt, Carleigh Samuels, Kelly Bono, Olivia Arter, Alyssa Valencia, Elianora Ovchiyan, Akshay Saluja, Mengesha Teshome, Alexandra Astarita, Sara Hubert, Tina Robinson, Mary G. Boyle, Alaina L. Robinson, Lisa M. Richardson, Tamair Curley, Jamie Mills, Elizabeth Capasso-Gulve, Sarah Pruitt-Welsh, Sam Kerz, Heena Kahn, Jamie Garner, Joshua Jackson, Wykita Willis-Lockhart, Michael Lehmkuhl, Vidhya Venkararaman

Vanderbilt University: Judy A. King, Erica Anderson, Brianna Schibley-Laird, Rebecca Tablada, Onika Abrams, Dandan Liu, Gaunchao Wang, Daniel Chandler, Gabriela Campos Umana, Sean Durkin, George “Preston” Gibson, Kaitlyn Menser, Juliet Umunna, Ivy Milligan, Catalina Padilla-Azain, Alexis Perry, Maria Paula Valencia Ortiz, Sarah Gaudieri, Krissy Paradissis, Suryakala Sarilla Bennet, Dayton Marchlewski

Columbia University: Liqun Wang, Anny L Diaz Perez, Raul A. Silverio Francisco, Ana Valdez de Romero

## Supplementary Materials

1. Text of daily diary questions related to medications and productivity losses; adapted influenza WPAI
2. Spreadsheet of per-dose OTC medication costs
3. Supplementary materials: tables, figures, and detailed methods for OTC medication doses and out-of-pocket costs

