## Supplemental methods and results for "Out-of-pocket costs and productivity losses associated with influenza illness and caregiving in a household study in the United States, 2024–2025"

**Supplementary tables and figures**

**Table S1.** Hourly productivity by age group (2025 USD)

| Age group | Market | Non-market |
|---|---|---|
| 15–24 years | $4.49 | $4.54 |
| 25–34 years | $18.10 | $10.86 |
| 35–44 years | $25.56 | $13.39 |
| 45–54 years | $27.37 | $9.94 |
| 55–64 years | $21.75 | $8.68 |
| 65–74 years | $7.78 | $9.45 |
| ≥75 years | $1.42 | $5.75 |

Note: Annual values from Gross et al. (2016) were converted to 2025 USD using the Cost Performance Index and then converted to daily values assuming a 365-day year and an 8-hour workday. Age-specific values were applied to each participant's hours of lost productivity to obtain per-illness costs.

Citation: Grosse SD, Krueger KV, Pike J. Estimated annual and lifetime labor productivity in the United States, 2016: implications for economic evaluations. J Med Econ. 2019;22(6):501-

**Tables S2–S5.** Percentage of participants who ever took OTC medications, duration of medication use, number of doses taken, and OTC medications costs (in 2025 USD) per illness

Note: Results are presented overall and by age group. Age 12 was used to separate age groups since that is the age at which OTC medications can generally be taken using adult dosing guidelines.
*n (%)
†Median and (IQR) among all participants with NMA influenza illness are reported except where indicated; means and bootstrapped 95% confidence intervals (CIs) for total days, doses, and costs of OTC medications are provided in the bottom row.

**Table S2.** Full results for medically attended influenza, maximum dose scenario (main analysis)

| | **Overall (N=594)** | | | | **Children aged <12 years (N=313)** | | | | **Adults and children aged 12–17 (N=281)** | | | |
|---|---|---|---|---|---|---|---|---|---|---|---|---|
| | Ever took* | Days† | Doses† | Cost† | Ever took* | Days† | Doses† | Cost† | Ever took* | Days† | Doses† | Cost† |
| Cold and flu or sinus medicine | 238 (40) | 0 (0–2) | 0 (0–4) | 0 (0–2.36) | 101 (32) | 0 (0–1) | 0 (0–2) | 0 (0–1.18) | 137 (49) | 0 (0–3) | 0 (0–6) | 0 (0–5.58) |
| Acetaminophen | 395 (66) | 2 (0–4) | 4 (0–8) | 1.16 (0–2.90) | 236 (75) | 3 (1–4) | 4 (0–8) | 2.32 (0–4.64) | 159 (57) | 1 (0–4) | 2 (0–7) | 0.38 (0–1.33) |
| Ibuprofen | 384 (65) | 2 (0–4) | 3 (0–7) | 1.08 (0–2.90) | 235 (75) | 2 (1–4) | 4 (0–8) | 2.32 (0–4.64) | 149 (53) | 1 (0–4) | 1 (0–6) | 0.18 (0–1.08) |
| Theraflu | 35 (6) | 0 (0–0) | 0 (0–0) | 0 (0–0) | 8 (3) | 0 (0–0) | 0 (0–0) | 0 (0–0) | 27 (10) | 0 (0–0) | 0 (0–0) | 0 (0–0) |
| Allergy medicine | 86 (14) | 0 (0–0) | 0 (0–0) | 0 (0–0) | 35 (11) | 0 (0–0) | 0 (0–0) | 0 (0–0) | 51 (18) | 0 (0–0) | 0 (0–0) | 0 (0–0) |
| Mucinex | 116 (20) | 0 (0–0) | 0 (0–0) | 0 (0–0) | 35 (11) | 0 (0–0) | 0 (0–0) | 0 (0–0) | 81 (29) | 0 (0–1) | 0 (0–1) | 0 (0–1.27) |
| Nasal spray | 55 (9) | 0 (0–0) | 0 (0–0) | 0 (0–0) | 17 (5) | 0 (0–0) | 0 (0–0) | 0 (0–0) | 38 (14) | 0 (0–0) | 0 (0–0) | 0 (0–0) |
| Sudafed | 26 (4) | 0 (0–0) | 0 (0–0) | 0 (0–0) | 4 (1) | 0 (0–0) | 0 (0–0) | 0 (0–0) | 22 (8) | 0 (0–0) | 0 (0–0) | 0 (0–0) |
| Naproxen | 14 (2) | 0 (0–0) | 0 (0–0) | 0 (0–0) | 5 (2) | 0 (0–0) | 0 (0–0) | 0 (0–0) | 9 (3) | 0 (0–0) | 0 (0–0) | 0 (0–0) |
| Zicam | 5 (1) | 0 (0–0) | 0 (0–0) | 0 (0–0) | 2 (1) | 0 (0–0) | 0 (0–0) | 0 (0–0) | 3 (1) | 0 (0–0) | 0 (0–0) | 0 (0–0) |
| Aspirin | 11 (2) | 0 (0–0) | 0 (0–0) | 0 (0–0) | 0 (0) | 0 (0–0) | 0 (0–0) | 0 (0–0) | 11 (4) | 0 (0–0) | 0 (0–0) | 0 (0–0) |
| Other medication | 23 (4) | 0 (0–0) | 0 (0–0) | 0 (0–0) | 4 (1) | 0 (0–0) | 0 (0–0) | 0 (0–0) | 19 (7) | 0 (0–0) | 0 (0–0) | 0 (0–0) |
| **All medications** | **572 (96)** | **5 (3–7)** | **12 (6–20)** | **6.38 (2.92–11.60)** | **306 (98)** | **5 (3–6)** | **11 (6–18)** | **6.50 (4.06–11.07)** | **266 (95)** | **5 (4–7)** | **13 (6–23)** | **6.03 (2.00–13.12)** |
| **Mean (95% CI)** | — | **5.14 (4.92, 5.32)** | **15.11 (14.12, 16.22)** | **9.11 (8.41, 10.09)** | — | **4.92 (4.66, 5.19)** | **13.16 (12.23, 14.24)** | **8.15 (7.47–9.02)** | — | **5.38 (5.02, 5.71)** | **17.32 (15.65, 19.43)** | **10.21 (8.76–11.85)** |

**Table S3.** Full results for non-medically attended influenza, maximum dose scenario (main analysis)

| | Overall (N=153) | | | | Children aged <12 years (N=38) | | | | Adults and children aged 12–17 (N=115) | | | |
|---|---|---|---|---|---|---|---|---|---|---|---|---|
| | Ever took* | Days† | Doses† | Cost† | Ever took* | Days† | Doses† | Cost† | Ever took* | Days† | Doses† | Cost† |
| Cold and flu or sinus medicine | 68 (44) | 0 (0–2) | 0 (0–4) | 0 (0–3.72) | 9 (24) | 0 (0–0) | 0 (0–0) | 0 (0–0) | 57 (50) | 0 (0–3) | 1 (0–6) | 0.93 (0–5.58) |
| Acetaminophen | 83 (54) | 1 (0–3) | 1 (0–5) | 0.38 (0–1.16) | 28 (74) | 1 (0.25–3) | 2 (0.25–5) | 1.16 (0–2.90) | 54 (47) | 0 (0–3) | 0 (0–4.5) | 0 (0–0.95) |
| Ibuprofen | 71 (46) | 0 (0–2) | 0 (0–4) | 0 (0–1.08) | 24 (63) | 1 (0–2) | 2 (0–4) | 1.16 (0–2.32) | 47 (41) | 0 (0–2) | 0 (0–2) | 0 (0–0.36) |
| Theraflu | 20 (13) | 0 (0–0) | 0 (0–0) | 0 (0–0) | 1 (2.6) | 0 (0–0) | 0 (0–0) | 0 (0–0) | 19 (17) | 0 (0–0) | 0 (0–0) | 0 (0–0) |
| Allergy medicine | 17 (11) | 0 (0–0) | 0 (0–0) | 0 (0–0) | 3 (7.9) | 0 (0–0) | 0 (0–0) | 0 (0–0) | 14 (12) | 0 (0–0) | 0 (0–0) | 0 (0–0) |
| Mucinex | 14 (9.2) | 0 (0–0) | 0 (0–0) | 0 (0–0) | 3 (7.9) | 0 (0–0) | 0 (0–0) | 0 (0–0) | 11 (9.6) | 0 (0–0) | 0 (0–0) | 0 (0–0) |
| Nasal spray | 6 (3.9) | 0 (0–0) | 0 (0–0) | 0 (0–0) | 0 (0) | 0 (0–0) | 0 (0–0) | 0 (0–0) | 6 (5.2) | 0 (0–0) | 0 (0–0) | 0 (0–0) |
| Sudafed | 5 (3.3) | 0 (0–0) | 0 (0–0) | 0 (0–0) | 0 (0) | 0 (0–0) | 0 (0–0) | 0 (0–0) | 5 (4.3) | 0 (0–0) | 0 (0–0) | 0 (0–0) |
| Naproxen | 2 (1.3) | 0 (0–0) | 0 (0–0) | 0 (0–0) | 0 (0) | 0 (0–0) | 0 (0–0) | 0 (0–0) | 2 (1.7) | 0 (0–0) | 0 (0–0) | 0 (0–0) |
| Zicam | 1 (0.7) | 0 (0–0) | 0 (0–0) | 0 (0–0) | 0 (0) | 0 (0–0) | 0 (0–0) | 0 (0–0) | 1 (0.9) | 0 (0–0) | 0 (0–0) | 0 (0–0) |
| Aspirin | 0 (0) | 0 (0–0) | 0 (0–0) | 0 (0–0) | 0 (0) | 0 (0–0) | 0 (0–0) | 0 (0–0) | 0 (0) | 0 (0–0) | 0 (0–0) | 0 (0–0) |
| Other medication | 3 (2.0) | 0 (0–0) | 0 (0–0) | 0 (0–0) | 1 (2.6) | 0 (0–0) | 0 (0–0) | 0 (0–0) | 2 (1.7) | 0 (0–0) | 0 (0–0) | 0 (0–0) |
| **All medications** | **137 (90)** | **3 (2–5)** | **7 (4–14)** | **3.72 (1.50–7.05)** | **36 (95)** | **3 (1.25–5)** | **7 (4.25–11.5)** | **4.60 (2.32–7.48)** | **101 (88)** | **3 (2–5)** | **8 (3–14.5)** | **3.20 (1.10–7.05)** |
| **Mean (95% CI)** | — | **3.38 (3.04, 3.74)** | **9.87 (8.36, 11.37)** | **6.03 (4.98, 7.32)** | — | **3.34 (2.68, 4.13)** | **8.74 (6.97, 11.02)** | **5.34 (4.29, 6.88)** | — | **3.39 (2.97, 3.77)** | **10.24 (8.67, 12.18)** | **6.26 (4.91, 8.14)** |

**Table S4.** Full results for medically attended influenza, minimum dose scenario (sensitivity analysis)

| | **Overall (N=594)** | | | | **Children aged <12 years (N=313)** | | | | **Adults and children aged 12–17 (N=281)** | | | |
|---|---|---|---|---|---|---|---|---|---|---|---|---|
| | Ever took* | Days† | Doses† | Cost† | Ever took* | Days† | Doses† | Cost† | Ever took* | Days† | Doses† | Cost† |
| Cold and flu or sinus medicine | 238 (40) | 0 (0–2) | 0 (0–3) | 0 (0–1.86) | 101 (32) | 0 (0–1) | 0 (0–2) | 0 (0–1.18) | 137 (49) | 0 (0–3) | 0 (0–5) | 0 (0–4.65) |
| Acetaminophen | 395 (66) | 2 (0–4) | 4 (0–8) | 1.14 (0–2.90) | 236 (75) | 3 (1–4) | 4 (0–8) | 2.32 (0–4.64) | 159 (57) | 1 (0–4) | 2 (0–7) | 0.38 (0–1.33) |
| Ibuprofen | 384 (65) | 2 (0–4) | 3 (0–6) | 1.08 (0–2.90) | 235 (75) | 2 (1–4) | 4 (0–7) | 2.32 (0–4.04) | 149 (53) | 1 (0–4) | 1 (0–6) | 0.18 (0–1.08) |
| Theraflu | 35 (6) | 0 (0–0) | 0 (0–0) | 0 (0–0) | 8 (3) | 0 (0–0) | 0 (0–0) | 0 (0–0) | 27 (10) | 0 (0–0) | 0 (0–0) | 0 (0–0) |
| Allergy medicine | 86 (14) | 0 (0–0) | 0 (0–0) | 0 (0–0) | 35 (11) | 0 (0–0) | 0 (0–0) | 0 (0–0) | 51 (18) | 0 (0–0) | 0 (0–0) | 0 (0–0) |
| Mucinex | 116 (20) | 0 (0–0) | 0 (0–0) | 0 (0–0) | 35 (11) | 0 (0–0) | 0 (0–0) | 0 (0–0) | 81 (29) | 0 (0–1) | 0 (0–1) | 0 (0–1.27) |
| Nasal spray | 55 (9) | 0 (0–0) | 0 (0–0) | 0 (0–0) | 17 (5) | 0 (0–0) | 0 (0–0) | 0 (0–0) | 38 (14) | 0 (0–0) | 0 (0–0) | 0 (0–0) |
| Sudafed | 26 (4) | 0 (0–0) | 0 (0–0) | 0 (0–0) | 4 (1) | 0 (0–0) | 0 (0–0) | 0 (0–0) | 22 (8) | 0 (0–0) | 0 (0–0) | 0 (0–0) |
| Naproxen | 14 (2) | 0 (0–0) | 0 (0–0) | 0 (0–0) | 5 (2) | 0 (0–0) | 0 (0–0) | 0 (0–0) | 9 (3) | 0 (0–0) | 0 (0–0) | 0 (0–0) |
| Zicam | 5 (1) | 0 (0–0) | 0 (0–0) | 0 (0–0) | 2 (1) | 0 (0–0) | 0 (0–0) | 0 (0–0) | 3 (1) | 0 (0–0) | 0 (0–0) | 0 (0–0) |
| Aspirin | 11 (2) | 0 (0–0) | 0 (0–0) | 0 (0–0) | 0 (0) | 0 (0–0) | 0 (0–0) | 0 (0–0) | 11 (4) | 0 (0–0) | 0 (0–0) | 0 (0–0) |
| Other medication | 23 (4) | 0 (0–0) | 0 (0–0) | 0 (0–0) | 4 (1) | 0 (0–0) | 0 (0–0) | 0 (0–0) | 19 (7) | 0 (0–0) | 0 (0–0) | 0 (0–0) |
| **All medications** | **572 (96)** | **5 (3–7)** | **11 (6–18)** | **5.80 (2.90–10.61)** | **306 (98)** | **5 (3–6)** | **10 (6–16)** | **5.92 (3.55–10.08)** | **266 (95)** | **5 (4–7)** | **12 (6–20)** | **5.58 (1.88–11.88)** |
| **Mean (95% CI)** | — | **5.14 (4.92, 5.32)** | **13.55 (12.72, 14.43)** | **7.98 (7.47, 8.66)** | — | **4.92 (4.66, 5.19)** | **12.21 (11.37, 13.21)** | **7.54 (7.01, 8.15)** | — | **5.38 (5.02, 5.71)** | **15.07 (13.53, 16.66)** | **8.47 (7.39, 9.54)** |

**Table S5.** Full results for non-medically attended influenza, minimum dose scenario (sensitivity analysis)

| | **Overall (N=153)** | | | | **Children aged <12 years (N=38)** | | | | **Adults and children aged 12–17 (N=115)** | | | |
|---|---|---|---|---|---|---|---|---|---|---|---|---|
| | Ever took* | Days† | Doses† | Cost† | Ever took* | Days† | Doses† | Cost† | Ever took* | Days† | Doses† | Cost† |
| Cold and flu or sinus medicine | 68 (44) | 0 (0–2) | 0 (0–3) | 0 (0–2.79) | 9 (24) | 0 (0–0) | 0 (0–0) | 0 (0–0) | 57 (50) | 0 (0–3) | 1 (0–4.5) | 0.93 (0–4.19) |
| Acetaminophen | 83 (54) | 1 (0–3) | 1 (0–5) | 0.38 (0–1.16) | 28 (74) | 1 (0.25–3) | 2 (0–5) | 1.16 (0–2.90) | 54 (47) | 0 (0–3) | 0 (0–5) | 0 (0–0.95) |
| Ibuprofen | 71 (46) | 0 (0–2) | 0 (0–4) | 0 (0–0.90) | 24 (63) | 1 (0–2) | 2 (0–4) | 1.16 (0–2.32) | 47 (41) | 0 (0–2) | 0 (0–2) | 0 (0–0.36) |
| Theraflu | 20 (13) | 0 (0–0) | 0 (0–0) | 0 (0–0) | 1 (2.6) | 0 (0–0) | 0 (0–0) | 0 (0–0) | 19 (17) | 0 (0–0) | 0 (0–0) | 0 (0–0) |
| Allergy medicine | 17 (11) | 0 (0–0) | 0 (0–0) | 0 (0–0) | 3 (7.9) | 0 (0–0) | 0 (0–0) | 0 (0–0) | 14 (12) | 0 (0–0) | 0 (0–0) | 0 (0–0) |
| Mucinex | 14 (9.2) | 0 (0–0) | 0 (0–0) | 0 (0–0) | 3 (7.9) | 0 (0–0) | 0 (0–0) | 0 (0–0) | 11 (9.6) | 0 (0–0) | 0 (0–0) | 0 (0–0) |
| Nasal spray | 6 (3.9) | 0 (0–0) | 0 (0–0) | 0 (0–0) | 0 (0) | 0 (0–0) | 0 (0–0) | 0 (0–0) | 6 (5.2) | 0 (0–0) | 0 (0–0) | 0 (0–0) |
| Sudafed | 5 (3.3) | 0 (0–0) | 0 (0–0) | 0 (0–0) | 0 (0) | 0 (0–0) | 0 (0–0) | 0 (0–0) | 5 (4.3) | 0 (0–0) | 0 (0–0) | 0 (0–0) |
| Naproxen | 2 (1.3) | 0 (0–0) | 0 (0–0) | 0 (0–0) | 0 (0) | 0 (0–0) | 0 (0–0) | 0 (0–0) | 2 (1.7) | 0 (0–0) | 0 (0–0) | 0 (0–0) |
| Zicam | 1 (0.7) | 0 (0–0) | 0 (0–0) | 0 (0–0) | 0 (0) | 0 (0–0) | 0 (0–0) | 0 (0–0) | 1 (0.9) | 0 (0–0) | 0 (0–0) | 0 (0–0) |
| Aspirin | 0 (0) | 0 (0–0) | 0 (0–0) | 0 (0–0) | 0 (0) | 0 (0–0) | 0 (0–0) | 0 (0–0) | 0 (0) | 0 (0–0) | 0 (0–0) | 0 (0–0) |
| Other medication | 3 (2.0) | 0 (0–0) | 0 (0–0) | 0 (0–0) | 1 (2.6) | 0 (0–0) | 0 (0–0) | 0 (0–0) | 2 (1.7) | 0 (0–0) | 0 (0–0) | 0 (0–0) |
| **All medications** | **137 (90)** | **3 (2–5)** | **7 (3–12)** | **3.41 (1.50–6.89)** | **36 (95)** | **3 (1.25–5)** | **6 (4–12)** | **4.31 (2.32–7.00)** | **101 (88)** | **3 (2–5)** | **7 (3–13)** | **3,13 (1.10–6.89)** |
| **Mean (95% CI)** | — | **3.38 (3.04, 3.74)** | **8.58 (7.62, 9.9)** | **5.01 (4.31, 6.02)** | — | **3.34 (2.68, 4.13)** | **7.89 (6.19, 9.68)** | **4.85 (3.95, 6.02)** | — | **3.39 (2.97, 3.77)** | **8.81 (7.51, 10.36)** | **5.07 (4.16, 6.41)** |

**Table S6.** Mean (95% CI) lost productivity hours and costs (in 2025 USD) due to illness, by age group and overall, among 747 participants with influenza illness.

| Age group | N ill | Hours missed from work/school | Market productivity loss (cost) | Hours missed from activities | Non-market productivity loss (cost) | Total hours missed | Total productivity loss (cost) |
|---|---|---|---|---|---|---|---|
| *Medically attended* | | | | | | | |
| 0–4 | 117 | 20.03 (16.53, 24.06) | — | 14.48 (11.28, 18.43) | — | 34.51 (27.8, 41.9) | — |
| 5–14 | 242 | 25.22 (23.35, 27.24) | — | 15.36 (13.66, 17.46) | — | 40.59 (37.14, 44.45) | — |
| 15–17 | 31 | 25.42 (20.84, 29.59) | 117.18 (98.74, 137.23) | 11.9 (8.48, 17.09) | 55.47 (39.86, 77.14) | 37.32 (30.48, 45.05) | 172.65 (143.46, 210.79) |
| 18–49 | 133 | 20.31 (17.88, 23.18) | 435.89 (374.08, 510.01) | 8.23 (6.36, 10.74) | 88.48 (67.44, 113.3) | 28.53 (24.92, 33.44) | 524.37 (443.44, 603.27) |
| ≥50 | 71 | 20.17 (16.59, 24.17) | 425.19 (333.63, 538.9) | 11.72 (8.72, 15.66) | 104.75 (78.69, 143.24) | 31.89 (27.32, 38.24) | 529.94 (430.47, 642.45) |
| All ages | 594 | 20.94 (18.98, 22.99) | 390.62 (343.53, 446.81) | 9.77 (8.24, 11.64) | 89.04 (75.44, 105.7) | 35.48 (33.32, 37.9) | 479.66 (429.17, 542.8) |
| *Non-medically attended* | | | | | | | |
| 0–4 | 21 | 12.61 (7.76, 19.91) | — | 3.57 (2.00, 6.19) | — | 16.19 (10.65, 22.91) | — |
| 5–14 | 24 | 14.5 (9.79, 20.72) | — | 10.13 (6.38, 17.04) | — | 24.625 (17.19, 38.47) | — |
| 15–17 | 7 | 16.0 (6.86, 30.29) | 73.76 (31.61, 147.52) | 12.57 (4.86, 30.86) | 58.58 (23.97, 146.68) | 28.57 (11.52, 60.71) | 132.34 (58.2, 271.1) |
| 18–49 | 86 | 13.36 (10.43, 16.92) | 313.49 (244.7, 398.0) | 8.71 (6.40, 12.11) | 103.72 (75.8, 141.4) | 22.07 (16.71, 28.77) | 417.21 (325.9, 519.1) |
| ≥50 | 15 | 13.20 (6.92, 19.22) | 289.06 (170.0, 435.7) | 9.00 (3.46, 18.40) | 82.22 (29.70, 153.51) | 22.20 (11.39, 37.08) | 371.28 (207.7, 552.8) |
| All ages | 153 | 13.54 (11.32, 15.87) | 294.56 (240.8, 364.4) | 8.43 (6.74, 10.82) | 97.80 (73.41, 130.99) | 21.97 (18.28, 25.96 | 392.36 (312.4, 495.9) |

Note: Bias-Corrected and Accelerated (BCa) confidence intervals were obtained using the "boot.ci" function in the *boot* package in R.

**Table S7.** Mean (95% CI) lost productivity hours and costs (in 2025 USD) associated with caregiving.

| Age of influenza case(s) in HH* | N HH | Any care-giving, n (%) | Hours missed from work by caregiver | Market productivity loss of caregiver (dollars) | Hours missed from activities by caregiver | Non-market productivity loss of caregiver (dollars) | Total hours missed by caregiver | Total productivity loss of caregiver (dollars) |
|---|---|---|---|---|---|---|---|---|
| Children only (aged <18 years) | | | | | | | | |
| 1 flu | 190 | 103 (54) | 9.54 (7.94, 11.45) | 220.89 (182.05, 266.23) | 6.25 (182.05, 266.23) | 78.29 (61.73, 99.91) | 15.79 (12.8, 19.11) | 299.18 (12.8, 19.11) |
| >1 flu | 44 | 30 (68) | 15.84 (11.09, 22.19) | 393.95 (285.94, 547.91) | 9.3 (5.83, 14.6) | 132.32 (85.28, 217.49) | 25.14 (17.62, 35.56) | 526.27 (375.77, 742.04) |
| Children only (<5 years) | | | | | | | | |
| 1 flu | 49 | 29 (59) | 9.55 (6.68, 13.22) | 193.5 (143.03, 299.73) | 6.94 (4.67, 10.94) | 85.34 (59.62, 146.95) | 16.49 (59.62, 146.95) | 278.84 (200.87, 416.32) |
| >1 flu | 6 | 4 (67) | 19.67 (3.22, 40.33) | 483.29 (72.78, 1011.12) | 12.17 (3.5, 31.71) | 217.87 (45.2, 635.72) | 31.83 (8.67, 64.33) | 701.17 (183.23, 1585.74) |
| Children only (5–17 years) | | | | | | | | |
| 1 flu | 141 | 74 (52) | 9.54 (7.57, 11.65) | 230.41 (183.23, 280.18) | 6.01 (4.45, 8.14) | 75.84 (57.25, 100.47) | 15.55 (12.42, 19.25) | 306.25 (249.69, 385.07) |
| >1 flu | 20 | 16 (80) | 17.2 (11.2, 26.8) | 460.08 (313.47, 717.48) | 8.5 (4.55, 21.02) | 120.83 (65.96, 275.65) | 25.7 (16.85, 44.22) | 580.91 (385.78, 928.32) |
| Adults only | | | | | | | | |
| 1 flu | 95 | 14 (15) | 1.43 (0.79, 2.64) | 21.5 (10.29, 46.81) | 1.29 (0.53, 3.15) | 13.36 (5.38, 33.79) | 2.73 (1.45, 5.34) | 34.86 (16.49, 72.78) |
| >1 flu | 34 | 11 (32) | 2.26 (1.12, 4.01) | 46.12 (20.76, 84.55) | 2.47 (0.59, 6.71) | 23.82 (7.43, 67.87) | 4.74 (2.15, 9.64) | 69.94 (36.32, 124.75) |
| Any age | | | | | | | | |
| 1 flu | 285 | 117 (41) | 6.84 (5.67, 8.07) | 154.43 (127.44, 186.41) | 11.44 (9.23, 14.04) | 56.65 (45.37, 72.63) | 11.44 (9.27, 13.74) | 211.08 (174.73, 256.49) |
| >1 flu | 192 | 122 (64) | 13.74 (11.56, 16.59) | 328.22 (271.33, 392.18) | 9.18 (7.24, 11.83) | 119.6 (93.25, 148.11) | 22.92 (18.64, 27.62) | 447.82 (380.61, 549.27) |

Note: Bias-Corrected and Accelerated (BCa) confidence intervals were obtained using the "boot.ci" function in the *boot* package in R.

### Supplementary Methods: Doses and costs of over-the-counter medications

#### 1. Creating the list of OTC medications

For this study, we developed a list of common OTC medications and medication categories to include in the 2024-25 RVTN daily diaries. We developed this list in an iterative process involving medical subject matter experts, study coordinators, and REDCap programmers. Considerations included the time and burden placed on the participants completing daily surveys; ensuring that the list would make sense to participants and reflect the most commonly taken OTC medications; and obtaining as much specificity as was feasible. We grouped generic and name brand medications together within each category, since participants may not distinguish between them and because the cost of OTC medications, even name brand ones, is generally low. The final list, as shown to participants, is provided below:

| |
|---|
| Cold and flu or sinus medicine *(Advil cold and sinus, Tylenol cold and flu, Dayquil, Nyquil, or store brand)* |
| Theraflu *(including store brand)* |
| Zicam *(including store brand)* |
| Ibuprofen *(Advil, Motrin, or store brand)* |
| Acetaminophen *(Tylenol, paracetamol, or store brand)* |
| Naproxen *(including Aleve or store brand)* |
| Aspirin *(including store brand)* |
| Mucinex *(including store brand)* |
| Nasal spray *(Flonase, Afrin, or store brand)* |
| Sudafed *(including store brand)* |
| Allergy medicines *(Zyrtec, Claritin, Xyzal, Allegra, Allerclear, Benadryl, or store brand)* |
| Other over the counter medication |

#### 2. Obtaining doses of OTC medications taken each day

When a participant responded "no" to having taken a medication on a specific day, the corresponding number of doses was analyzed as 0.

When a participant responded "yes" to having taken a medication, they were then asked to select the number of doses from a dropdown list with choices 1, 2, or 3+ doses. The question was asked this way to reduce the burden on participants while promoting data quality; since most OTC medications are only taken a few times per day, these answer choices are the most likely responses. If a participant chose 1 or 2, the number of doses was analyzed as 1 or 2, respectively.

Participants chose the "3+" response for 97 out of 737 (13%) total medication instances, most commonly for "cold and flu or sinus" medication. To convert the categorical "3+" response to a numeric one for analysis, we considered two scenarios: the "minimum scenario" in which 3+ was analyzed as "exactly 3" doses, and a "maximum scenario" in which 3+ was analyzed as the maximum plausible daily dose based on medication guidelines (Table xx). We considered minimum and maximum scenarios for children aged <12 years and for adults and children aged ≥ 12 years, as this is the age at which medications can typically be dosed the same as for adults. Results for the maximum scenario are presented in the main text, and results for the minimum scenario are presented below.

**Adults and children aged ≥12 years** (the age when OTC medications can typically be dosed the same as for adults):

| **Medication** | **Dosing assumptions*** | **Minimum Scenario** | **Maximum Scenario** |
|---|---|---|---|
| Cold and flu or sinus meds | Depends on product; usually taken every 4-6 hours for a max of 6 doses | 3 | 6 |
| Theraflu | "Do not exceed 3 packets per day" | 3 | 3 |
| Zicam | Nasal: 4-6 doses per day; lozenges: 6 doses | 3 | 6 |
| Ibuprofen | Every 6 hours for 200 mg tablets | 3 | 4 |
| Acetaminophen | Every 6 hours for 500 mg tablets, up to 3,000 mg/day | 3 | 4 |
| Naproxen | Every 8-12 hours, up to 660 mg/day | 3 | 3 |
| Aspirin | Every 4-6 hours, up to 4,000 mg/day | 3 | 6 |
| Mucinex | Assuming regular Mucinex: every 12 hours, so the max should be 2. If 3+ then it might be "immediate-release guaifenesin 200 mg" which has a max of 6/day | 3 | 6 |
| Nasal spray | Afrin: max 2 times per day<br>Regular steroid spray: 1 time per day<br>Saline: unlimited use | 3 | 6 |
| Sudafed | 60 mg every 4-6 hours, max 4 times per day | 3 | 4 |
| Allergy medicines | If they're taking more than 1 dose per day it's probably Benadryl. Max 4 | 3 | 4 |
| Other | -- | 3 | 6 |

**Children aged <12 years:**

| **Medication** | **Dosing assumptions*** | **Minimum Scenario** | **Maximum Scenario** |
|---|---|---|---|
| Cold and flu or sinus meds | **Not recommended under age 4**<br>Ages **4–6: only if clinician advises**<br>Ages **6–12: usually every 4–6 hours**<br>**Max: 4 doses per day** | 3 | 4 |
| Theraflu | Not recommended under 12 | NA | NA |
| Zicam | Not recommended under 12 | NA | NA |
| Ibuprofen | **For ages ≥ 6 months only**<br>**Dosing:** 10 mg/kg<br>**Frequency: Every 6–8 hours**<br>**Maximum: 4 doses in 24 hours** | 3 | 4 |
| Acetaminophen | **Dosing:** 10–15 mg/kg<br>**Frequency: Every 4–6 hours**<br>**Maximum: 5 doses in 24 hours** | 3 | 5 |
| Naproxen | Not recommended under 12 | NA | NA |
| Aspirin | Not recommended under 18 | NA | NA |

| | | | |
|---|---|---|---|
| Mucinex | **Immediate Release Liquid / Chewables**<br>• Usually **every 4 hours**<br>• **Max 6 doses/day** | 3 | 6 |
| Nasal spray | Afrin: max 2 times per day<br>Regular steroid spray: 1 time per day<br>Saline: unlimited use | 3 | 6 |
| Sudafed | Ages 6+: 60 mg every 4-6 hours, max 4 times per day | 3 | 4 |
| Allergy medicines | 1/day (Zyrtec & Claritin); 1-2/day depending on strength (Allegra); up to 4/day (Benadryl) | 3 | 4 |
| Other | -- | 3 | 6 |

#### 3. Imputing missing doses

We imputed missing data when a participant said "yes" to taking a medication on a specific day but then left the corresponding number of doses blank (i.e., did not select 1, 2, or 3+ doses). The number of doses was missing for 24 out of 737 (3%) medication instances. We imputed using a person-specific approach first, followed by a medication-specific approach if needed.

- *Person-specific:* if the participant had given a dosage of that medication on other day(s), we used the median number of doses for that medication and person
- *Medication-specific:* if the participant did not give a dosage of that medication on any other day, we used the median number of doses for that medication across all participants (stratified by age <12 vs. 12+ years)

#### 4. Obtaining per-dose costs of OTC medications

We used online retail websites to obtain 2025 prices for OTC medications. Since each medication category in our analysis represented many possible products, we used the following approach to obtain per-dose costs:

- First, we selected 2-5 representative products per category. We included both generic and name brand, as applicable, and used mid-size containers to not underestimate or overestimate the per-dose cost (medications bought in bulk would be less expensive per dose, while travel size medications would be more expensive per dose)
- Next, we obtained costs from retail websites for a variety of sources: a retail pharmacy (CVS), a big box store (Walmart), and as online retailer (Amazon). We did not use [databases] because they usually reflect the wholesale price available to stores, whereas our analysis focused on the out-of-pocket cost to the consumer.
- These costs are documented in the supplementary tables below.

**Supplementary Methods for per-dose costs of OTC medications**

1. Created a list of representative products
   - Used a mix of name-brand and generic products
   - Aimed for mid-sized packaging to avoid over- or under-estimating the per-dose price
   - Aimed to have the exact same products/sizes represented across all sources, but if that was not possible, we chose the most popular products based on number of reviews and/or "overall pick"
   - Some products (e.g. "real" Sudafed) were not available from all sources

2. Sources
   - Used websites for a variety of settings
   - We used non-sale, national, online prices
   - Retail pharmacy: CVS (generics = CVS brand)
   - Bog box store: Walmart (generics = Equate)
   - Online retailer: Amazon (generics = Amazon brand)

3. Dosing
   - Used standard dosing guidelines to get per-dose price.
   - If the guideline gave options (e.g., 1-2 pills, 5-10 mL, 1-2 sprays), we chose the higher option.
   - For kids, we used an intermediate dose (e.g. 10 mL if dose ranged from 5-15 mL based on age/weight).

**Children <12 years**

| Medication | Product | Retail pharmacy | | | Big box store | | | Online retailer | | | Average $/dose | Dosing guidelines |
|---|---|---|---|---|---|---|---|---|---|---|---|---|
| | | $/product | $/pill | $/dose | $/product | $/pill | $/dose | $/product | $/pill | $/dose | | |
| **Cold and flu or sinus meds** | DayQuil kids | 14.29 | 0.06 | 0.91 | 9.97 | 0.04 | 0.63 | 18.99 | 0.04 | 0.60 | 0.59 | 15 mL |
| | Children's tylenol cold & flu | 10.99 | 0.09 | 0.92 | 7.48 | 0.06 | 0.63 | 7.12 | 0.06 | 0.60 | | Assume 10 mL dose |
| | Children's Robitussin | 9.99 | 0.08 | 0.42 | 6.48 | 0.05 | 0.27 | 8.99 | 0.08 | 0.38 | | Assume 5 mL dose (children 6-11 can take 10) |
| | Children's Dimetapp | 8.99 | 0.08 | 0.76 | 6.27 | 0.05 | 0.53 | 9.96 | 0.04 | 0.42 | | 10 mL dose (only for children 6-11) |
| **Theraflu** | | | | | | | | | | | 1.15 | Not recommended for children <12 |
| **Zicam** | | | | | | | | | | | 0.86 | Not recommended for children <12 |
| **Ibuprofen** | Children's advil (liquid) | 8.49 | 0.07 | 0.71 | 6.18 | 0.05 | 0.52 | 7.39 | 0.06 | 0.63 | 0.58 | Used 10 mL dose (median dose by weight) |
| | Generic children's ibuprofen (liquid) | 7.59 | 0.06 | 0.64 | 4.16 | 0.04 | 0.35 | 6.52 | 0.03 | 0.28 | | Used 10 mL dose (median dose by weight) |
| | Children's motrin (liquid) | 9.49 | 0.08 | 0.79 | 7.68 | 0.07 | 0.65 | 7.68 | 0.07 | 0.65 | | Used 10 mL dose (median dose by weight) |

| | | | | | | | | | | | | |
|---|---|---|---|---|---|---|---|---|---|---|---|---|
| **Acetaminophen** | Children's tylenol (liquid) | 9.29 | 0.08 | 0.77 | 7.68 | 0.07 | 0.65 | 7.68 | 0.07 | 0.65 | 0.56 | Used 10 mL dose (median dose by weight) |
| | Generic children's acetaminophen (liquid) | 7.49 | 0.06 | 0.63 | 3.48 | 0.03 | 0.29 | 8.50 | 0.04 | 0.36 | | Used 10 mL dose (median dose by weight) |
| **Naproxen** | | | | | | | | | | | 0.15 | Not recommended for children <12 |
| **Aspirin** | | | | | | | | | | | 0.12 | Not recommended for children <12 |
| **Mucinex** | Mucinex children's mighty chews | 18.99 | 1.19 | 2.37 | 13.97 | 0.87 | 1.75 | 13.97 | 0.87 | 1.75 | 1.23 | |
| | Mucinex children's liquid | 16.49 | 0.14 | 0.70 | 9.97 | 0.08 | 0.42 | 9.97 | 0.08 | 0.42 | | used 5 mL dose (but children 6-11 can take 10) |
| **Nasal spray** | Generic children's saline nasal spray | 4.79 | 0.12 | 0.24 | | | | | | | 0.61 | 1 spray per nostril (2 sprays per dose) |
| | Children's flonase | 20.49 | 0.28 | 1.14 | 21.99 | 0.31 | 1.22 | 12.99 | 0.18 | 0.18 | | 2 sprays per nostril (4 sprays per dose) |
| | Afrin Children's Saline spray | 12.99 | 0.13 | 0.26 | | | | | | | | 1 spray per nostril (2 sprays per dose) |
| **Sudafed** | Children's Sudafed PE | 11.49 | 0.10 | 0.49 | 6.87 | 0.06 | 0.29 | 13.38 | 0.11 | 0.57 | 0.46 | used 5 mL dose (but |

| | | | | | | | | | | | | |
|---|---|---|---|---|---|---|---|---|---|---|---|---|
| | Nasal Decongestant (liquid) | | | | | | | | | | | children 6-11 can take 10) |
| | Children’s Sudafed Nasal Decongestant (liquid) | 11.49 | 0.10 | 0.49 | | | | | | | | used 5 mL dose (but children 6-11 can take 10) |
| **Allergy meds** | Children's Claritin Chewables | 26.59 | 0.89 | 0.89 | 24.97 | 0.83 | 0.83 | 24.32 | 0.61 | 0.61 | 0.76 | |
| | Children's Zyrtec | 25.79 | 1.07 | 1.07 | 19.96 | 0.83 | 0.83 | 13.44 | 0.11 | 1.14 | | |
| | Children's Benadryl | 13.49 | 0.06 | 0.57 | 10.44 | 0.04 | 0.44 | 10.69 | 0.05 | 0.45 | | used 10 mL (dose is "5-10" mL) and only 6+ |
| **Other** | (Average of all the above) | | | | | | | | | | 0.64 | |

### Ages 12 and older

| Medication | Product | Retail pharmacy | | | Big box store | | | Online retailer | | | Average $/dose | Dosing guidelines |
|---|---|---|---|---|---|---|---|---|---|---|---|---|
| | | $/product | $/pill | $/dose | $/product | $/pill | $/dose | $/product | $/pill | $/dose | | |
| **Cold and flu or sinus meds** | DayQuil cold & flu | 27.49 | 0.57 | 1.15 | 22.93 | 0.48 | 0.96 | 19.92 | 0.415 | 0.83 | 0.93 | max 8 caplets/24 h, 2 caps/4 h |
| | Tylenol cold & flu | 10.79 | 0.90 | 1.80 | 7.67 | 0.32 | 0.64 | 8.99 | 0.37 | 0.75 | | max 10 caplets/24 h (2 every 4 h) |
| | Alka-Seltzer plus cold & flu | 7.99 | 0.67 | 1.33 | 9.47 | 0.39 | 0.79 | 8.89 | 0.37 | 0.74 | | max 8 tabs/24 h, 2 tabs/4 h |
| | Generic cold & flu medication | 10.79 | 0.67 | 1.35 | 4.98 | 0.21 | 0.42 | 10.49 | 0.22 | 0.44 | | max 8 caplets/24 h, 2 caps/4 h |
| **Theraflu** | Theraflu Daytime Severe Cold & Cough (packets) | 7.99 | 1.33 | 1.33 | 7.98 | 1.33 | 1.33 | 7.98 | 1.33 | 1.33 | 1.15 | 1 packet/6 hours; max 3 per day |
| | Theraflu Nighttime Severe Cold & Cough (packets) | 7.99 | 1.33 | 1.33 | 7.98 | 1.33 | 1.33 | 7.98 | 1.33 | 1.33 | | 1 packet/6 hours; max 3 per day |
| | Theraflu Day and Night caplets | 17.49 | 0.44 | 0.87 | 12.98 | 0.32 | 0.65 | 17.49 | 0.44 | 0.87 | | 2 caplets/6 hours; max 6 caplets per day (3 doses) |
| **Zicam** | Zicam Cold Remedy RapidMelts, 25 count | 14.49 | 0.58 | 0.58 | 10.98 | 0.44 | 0.44 | 10.98 | 0.44 | 0.44 | 0.86 | 1 per 3 hours |
| | Zicam Cold Remedy Medicated Lozenges, 25 count | 14.49 | 0.58 | 0.58 | 16.66 | 0.67 | 0.67 | 9.99 | 0.40 | 0.40 | | |

| | | | | | | | | | | | | |
|---|---|---|---|---|---|---|---|---|---|---|---|---|
| | Zicam Cold Remedy Nasal Spray | 17.99 | 0.45 | 2.70 | 10.98 | 0.27 | 1.10 | 8.17 | 0.20 | 0.82 | | CVS: ultra spray: 2-3 sprays per nostril/12 hours/max 2 doses per day; Walmart & Amazon: regular Zicam spray--2 sprays per nostril/3 hours/max 5 doses (all: assume 40 sprays per 0.5 fl oz bottle) |
| **Ibuprofen** | Generic ibuprofen, 200 mg, 100 count | 9.79 | 0.10 | 0.20 | 1.98 | 0.02 | 0.04 | 2.33 | 0.02 | 0.05 | 0.18 | |
| | Advil, 200 mg, 100 count | 12.99 | 0.13 | 0.26 | 10.98 | 0.11 | 0.22 | 10.98 | 0.11 | 0.22 | | |
| | Motrin, 200 mg, 100 count | 12.49 | 0.12 | 0.25 | 9.47 | 0.09 | 0.19 | 9.47 | 0.09 | 0.19 | | |
| **Acetaminophen** | Generic acetaminophen, 500 mg, 150/200 count | 11.99 | 0.08 | 0.16 | 3.94 | 0.02 | 0.04 | 4.06 | 0.04 | 0.08 | 0.19 | 2 pills/4-6 hours, max 6 pills per day |
| | Tylenol, 375 mg, 100 count | 12.49 | 0.12 | 0.25 | 6.96 | 0.07 | 0.14 | 6.96 | 0.07 | 0.14 | | 2 pills/4-6 hours, max 10 pills per day |
| | Tylenol extra strength, 500 mg, 25/100/100 count | 5.99 | 0.24 | 0.48 | 10.97 | 0.11 | 0.22 | 12.04 | 0.12 | 0.24 | | 2 pills/4-6 hours, max 6 pills per day |
| **Naproxen** | Generic naproxen, 220 mg | 12.99 | 0.13 | 0.13 | 5.17 | 0.06 | 0.06 | 6.38 | 0.07 | 0.07 | 0.15 | 1/8-12 hours, max 3 per day |
| | Aleve, 220 mg | 16.79 | 0.19 | 0.19 | 11.67 | 0.13 | 0.13 | 11.99 | 0.13 | 0.13 | | |

| | | | | | | | | | | | |
|---|---|---|---|---|---|---|---|---|---|---|---|
| | Aleve liquid gels, 220 mg | 18.79 | 0.23 | 0.23 | 13.97 | 0.17 | 0.17 | 28.78 | 0.24 | 0.24 | | |
| **Aspirin** | Bayer aspirin | 9.29 | 0.09 | 0.19 | 6.97 | 0.07 | 0.14 | 10.97 | 0.05 | 0.11 | 0.12 | 2 pills per dose |
| | Generic aspirin | 8.29 | 0.08 | 0.17 | 1.37 | 0.01 | 0.03 | 5.99 | 0.05 | 0.10 | | 2 pills per dose |
| **Mucinex** | Mucinex Fast-Max Cold & Flu / Cold, Flu & Sore Throat | 23.49 | 0.98 | 1.96 | 18.47 | 0.77 | 1.54 | 18.47 | 0.77 | 1.54 | 1.27 | max 12 gels/24 hours; 2 gels every 6 h |
| | Mucinex 12-hour (regular strength) | 30.99 | 0.77 | 1.55 | 14.97 | 0.75 | 1.50 | 11.00 | 0.55 | 1.10 | | max 4 tabs/24 hours; 1-2 tabs every 12 hours |
| | Mucinex D/DM (regular strength) | 33.79 | 0.84 | 1.69 | 14.97 | 0.75 | 1.50 | 12.08 | 0.60 | 1.21 | | max 4 tabs/24 hours; 1-2 tabs every 12 hours |
| | Generic guaifenesin maximum strength (1200 mg) | 32.99 | 0.79 | 0.79 | 16.32 | 0.58 | 0.58 | 19.99 | 0.29 | 0.29 | | 1 tab every 12 hours |
| **Nasal spray** | Afrin Original | 17.49 | 0.06 | 0.35 | 7.47 | 0.05 | 0.30 | 7.47 | 0.05 | 0.20 | 0.40 | 2-3 sprays/nostril/2x per day (up to 12 sprays/day) |
| | Afrin store brand (oxymetazoline 0.05%) | 12.49 | 0.04 | 0.25 | 4.88 | 0.02 | 0.10 | 4.19 | 0.01 | 0.06 | | 2-3 sprays/nostril/2x per day (up to 12 sprays/day) |
| | Flonase | 31.49 | 0.22 | 0.87 | 17.48 | 0.24 | 0.97 | 17.48 | 0.24 | 0.97 | | 2 sprays/nostril/2x per day (8 sprays/day) |
| | Generic flonase | 14.49 | 0.20 | 0.81 | 9.87 | 0.14 | 0.55 | 13.11 | 0.09 | 0.36 | | 2 sprays/nostril/2x per day (8 sprays/day) |
| | Saline spray | 5.49 | 0.02 | 0.09 | 1.97 | 0.01 | 0.02 | 2.75 | 0.01 | 0.03 | | Unlimited |

| | | | | | | | | | | | | |
|---|---|---|---|---|---|---|---|---|---|---|---|---|
| **Sudafed** | Sudafed, 30 mg | 16.79 | 0.35 | 0.70 | 13.86 | 0.29 | 0.58 | | | | 0.71 | max 240 mg/day (2 pills x 4 times per day) |
| | Generic sudafed 30 mg | 12.99 | 0.27 | 0.54 | 4.48 | 0.19 | 0.37 | | | | | " |
| | Sudafed 12-hour, 120 mg | 16.79 | 0.84 | 0.84 | | | | | | | | max: 240 mg/day (take 2x per day) |
| | Generic sudafed 12-hour | 13.79 | 0.69 | 0.69 | 6.98 | 0.35 | 0.35 | | | | | " |
| | Sudafed PE (sinus congestion, head congestion) | 11.79 | 0.49 | 0.98 | 14.51 | 0.60 | 1.21 | | | | | max: 60 mg/day (10 mg x every 6h) |
| | Generic sudafed PE | 8.99 | 0.37 | 0.75 | | | | 8.99 | 0.37 | 0.75 | | " |
| **Allergy meds** | Claritin | 22.49 | 1.12 | 1.12 | 19.97 | 0.67 | 0.67 | 18.15 | 0.61 | 0.61 | 0.67 | |
| | Generic claritin (loratidine) | 16.79 | 0.84 | 0.84 | 7.88 | 0.20 | 0.20 | 4.53 | 0.15 | 0.15 | | |
| | Allegra | 18.49 | 1.23 | 1.23 | 19.96 | 0.67 | 0.67 | 13.99 | 0.58 | 0.58 | | |
| **Other** | (Average of all the above) | | | | | | | | | | 0.60 | |
