## Supplementary material for "Out-of-pocket costs and productivity losses associated with influenza illness and caregiving in a household study in the United States, 2024–2025": Text of questionnaires

**Productivity loss questionnaires**

**Daily diaries:**

- Administered daily to all participants (parents completed on behalf of children)
- Questions about productivity losses from work, school, and other activities, due to own illness vs. caretaking for another ill household member

| **Next, we will ask you some questions about activities.** | |
|---|---|
| On [calendar date], did [participant name] have to do any of the following? | |
| Stay home from school, work, or miss other activities because they were sick? | □ Yes*<br>□ No<br>□ Not sure<br>□ Prefer not to answer |
| *If yes, approximately how many hours of school or work did [participant name] miss due to illness? | [free-text field with integer validation and max of 24] |
| *If yes, approximately how many hours of activities, other than work or school, did [participant name] miss due to illness? | [free-text field with integer validation and max of 24] |
| Stay home from school, work, or miss other activities because someone else in the household was sick and they needed to care for them? | □ Yes*<br>□ No<br>□ Not sure<br>□ Prefer not to answer |
| *If yes, approximately how many hours of school or work did [participant name] miss to care for someone else in the household who was sick? | [free-text field with integer validation and max of 24] |
| *If yes, approximately how many hours of activities, other than work or school, did [participant name] miss to care for someone else in the household who was sick? | [free-text field with integer validation and max of 24] |
| Stay home from school, work, or miss other out of the house activities because someone else in the household was sick and they were concerned that they might also get sick? | □ Yes<br>□ No<br>□ Not sure<br>□ Prefer not to answer |

**30-day follow up:** modified Work Productivity and Activity Impairment (WPAI) questionnaire

- Administered only to adults
- WPAI is a validated survey instrument that we modified to
  - quantify both market (work) and non-market (other activities) productivity losses
  - quantify productivity losses due to own illness vs. caretaking for another ill household member

| | |
|---|---|
| ***[Questions for all adult Participants]***<br>**Please answer the following questions about the impact of illness in your household on your ability to work.** | |
| 1. During the first 4 weeks of influenza illness in your household, who had influenza? (select all that apply) | □ Myself<br>□ Other household member(s)<br>□ Prefer not to answer |
| 2. Are you currently employed (working for pay)? | □ No *(skip to page 5)*<br>□ Yes *(continue)*<br>□ Prefer not to answer *(skip to page 5)* |
| **If you answered "yes" to question 2, please answer the following questions about your job and the impact of influenza illness in your household on your ability to work.** | |
| 3. During the first four weeks of influenza illness in your household, were you able to work remotely? | □ No<br>□ Yes<br>□ Prefer not to answer |
| 4. If you miss work because you are sick or because you need to care for someone who is sick, do you still get paid?<br>*(i.e., do you have a certain amount of paid sick time that you can use each year?)* | □ No<br>□ Yes<br>□ I don't know<br>□ Prefer not to answer |
| 5. If you had influenza, did you miss time from work because of your illness? | □ No<br>□ Yes *(complete the question below)*<br>□ Prefer not to answer |

*If yes,* During the first four weeks of influenza illness in your household, how many hours did you miss work because of problems related to **your** influenza illness? Please include hours you missed for sick days, times you went in late or left early, and any other time you missed from work because of **your** illness. *As a reminder, a typical workday for a full time worker in the US is 8 hours and a typical work week is 40 hours.*

| **Week:** | **Number of hours missed from work because of your illness** |
|---|---|
| **1** | |
| **2** | |
| **3** | |
| **4** | |

| | |
|---|---|
| 6. If another household member had influenza, did you miss time from work in order to care for others in your household with the flu? | □ No<br>□ Yes *(complete the question below)*<br>□ Prefer not to answer |

*If yes,* During the first four weeks of influenza illness in your household, how many hours did you miss work because of problems related to **another person's** influenza illness (e.g., taking care of them? Please include all hours you missed because of **another person's** illness.

| Week: | Number of hours missed from work because of a household member's illness |
|---|---|
| 1 | |
| 2 | |
| 3 | |
| 4 | |

7. During the first four weeks influenza illness in your household, how many hours did you miss work because of any other reason other than illness? This would include vacation, holidays, or other time off?

*As a reminder, a typical workday for a full time worker in the US is 8 hours and a typical work week is 40 hours.*

| Week: | Number of hours missed from work not because of illness |
|---|---|
| 1 | |
| 2 | |
| 3 | |
| 4 | |

8. During the first four weeks of influenza illness in your household, how many hours did you actually work?

*As a reminder, a typical workday for a full time worker in the US is 8 hours and a typical work week is 40 hours.*

| Week | Hours worked (*It's okay to give your best estimate*) | Was this a typical work week for you, in terms of the number of hours you worked? |
|---|---|---|
| 1 | | □ Yes □ No □ Prefer not to answer |
| 2 | | □ Yes □ No □ Prefer not to answer |
| 3 | | □ Yes □ No □ Prefer not to answer |
| 4 | | □ Yes □ No □ Prefer not to answer |

9. During the first 4 weeks of influenza illness in your household, how much did this illness affect your productivity while you were working? Please estimate how much influenza illness in your household affected your productivity <u>while you were working</u> for each week using a scale from 0 (no effect on your work) to 10 (completely prevented from working).

*Think about days that you were limited in the amount or kind of work that you could do, days when you accomplished less than you would have liked, or days when you could not do you work as well or as carefully as usual. If influenza illness only affected your work a little, choose a low number. If influenza illness affected your work a lot, choose a high number.*

| Week | No effect on work 0 | 1 | 2 | 3 | 4 | 5 | 6 | 7 | 8 | 9 | Completely prevented from working 10 |
|---|---|---|---|---|---|---|---|---|---|---|---|
| 1 | □ | □ | □ | □ | □ | □ | □ | □ | □ | □ | □ |
| 2 | □ | □ | □ | □ | □ | □ | □ | □ | □ | □ | □ |
| 3 | □ | □ | □ | □ | □ | □ | □ | □ | □ | □ | □ |
| 4 | □ | □ | □ | □ | □ | □ | □ | □ | □ | □ | □ |

| | |
|---|---|
| ***[Questions for all adult Participants]***<br>**Please answer the following questions about the impact of influenza illness on <u>your</u> ability to perform your regular activities.**<br>*By regular activities, we mean the usual activities you do, such as work around the house, shopping, childcare, exercising, studying, etc.* | |
| 10. During the first 4 weeks of influenza illness in your household, who had influenza? (select all that apply) | □ Myself *(answer question 10a)*<br>□ Other household member(s) *(answer question 10b)*<br>□ Prefer not to answer *(skip to question 11)* |
| 10a. Did you miss time from activities other than work because you had influenza? | □ No<br>□ Yes *(complete the below question)*<br>□ Prefer not to answer |

*If yes,* During the first four weeks of influenza illness in your household, how many hours did you miss from your regular daily activities, other than work at a job, because of problems related to **your** influenza illness? Please include hours you missed because of **your** illness.

| **Week:** | **Number of hours missed from regular daily activities because of your illness** |
|---|---|
| **1** | |
| **2** | |
| **3** | |
| **4** | |

| | |
|---|---|
| 10b. Did you miss time from activities other than work in order to care for others in your household with the flu? | □ No<br>□ Yes *(complete the below question)*<br>□ Prefer not to answer |

*If yes,* During the first four weeks of influenza illness in your household, how many hours did you miss from your regular daily activities, other than work at a job, because of problems related to **another person's** influenza illness (e.g., taking care of them)? Please include all hours you missed because of **another person's** illness.

| **Week:** | **Number of hours missed from regular daily activities because of a household member's illness** |
|---|---|
| **1** | |
| **2** | |
| **3** | |
| **4** | |
